# Trends in Medicare Part D Spending on Antifibrotic Therapies for Pulmonary Fibrosis: 2019–2023

**DOI:** 10.1101/2025.08.01.25332591

**Authors:** Shaheen Rizly, Divya Vaithiswaran, Adrian Lorenzana, Anthony Saleh

## Abstract

**Background:** Nintedanib and pirfenidone are antifibrotic agents approved for the treatment of idiopathic pulmonary fibrosis (IPF) and progressive fibrosing interstitial lung diseases. Despite their clinical significance, national trends in Medicare Part D spending and utilization for these therapies remain insufficiently characterized.

**Objective:** To evaluate national trends in Medicare Part D spending, claims, and cost-per-claim for nintedanib and pirfenidone from 2019 to 2023.

**Methods:** Medicare Part D Drug Spending Dashboard data were analyzed for total spending, number of claims, and average spending per claim for nintedanib and pirfenidone. Trends over five years were assessed.

**Results:** Total spending on nintedanib increased from 2019 to 2023, with a consistent rise in both claims and average cost per claim. In contrast, pirfenidone’s total spending declined sharply, primarily due to a reduction in claims. The average cost per claim for both drugs increased over the study period.

**Conclusion:** Medicare spending on antifibrotic therapies is increasingly dominated by nintedanib, reflecting evolving prescribing patterns and potential differences in tolerability or access. These findings have implications for cost-containment strategies and formulary management.

**Author Summary:** Idiopathic pulmonary fibrosis (IPF) is a progressive lung disease treated with two antifibrotic medications: nintedanib and pirfenidone. These therapies are costly, and little is known about how Medicare has been spending on them in recent years. Using publicly available Medicare data from 2019 to 2023, we found that spending on nintedanib has increased significantly, while spending on pirfenidone has dropped. The average cost per prescription rose for both drugs. These findings may reflect changes in prescribing habits, patient access, and insurance coverage, and highlight the need to monitor drug costs and usage to ensure affordable treatment for patients.

## Introduction

Idiopathic pulmonary fibrosis (IPF) is a progressive, fatal interstitial lung disease characterized by irreversible fibrosis and declining lung function, with a median survival of less than five years from diagnosis ^1^. The American Thoracic Society, in collaboration with the European Respiratory Society, Japanese Respiratory Society, and Latin American Thoracic Association, recommends antifibrotic therapy with nintedanib or pirfenidone as standard of care for IPF and select cases of progressive fibrosing interstitial lung diseases (PF-ILDs) ^2^. Both agents have demonstrated efficacy in slowing the rate of forced vital capacity (FVC) decline and reducing the risk of acute exacerbations, though neither halts or reverses disease progression ^3–6^.Nintedanib, a tyrosine kinase inhibitor, and pirfenidone, a pyridone derivative, are associated with distinct adverse event profiles, most commonly gastrointestinal symptoms for nintedanib and photosensitivity or rash for pirfenidone ^7–8^.

The introduction of antifibrotic therapies has redefined the management of IPF and PF-ILDs, but their high acquisition costs have raised concerns regarding economic sustainability and access, particularly within the Medicare population ^9–13^. While both drugs are effective in slowing disease progression, adoption rates remain low, with only a minority of eligible patients initiating therapy since approval ^9^. High out-of-pocket costs and drug tolerability are significant barriers to broader adoption, and discontinuation rates are substantial for both agents ^9^. Comparative analyses suggest that while clinical outcomes such as mortality and hospitalization are similar between nintedanib and pirfenidone, drug costs and persistence may differ, with pirfenidone associated with lower respiratory-related costs and longer time to discontinuation in some cohorts ^10, 11^. From an economic perspective, annual drug costs for both agents are high, and total healthcare expenditures for patients with IPF are significantly increased following initiation of antifibrotic therapy ^10, 11^. However, treatment is associated with reduced mortality and hospitalization rates among Medicare beneficiaries, supporting the value of antifibrotic therapy despite the high upfront costs ^12^.The American Thoracic Society guideline emphasizes the importance of early diagnosis and timely initiation of antifibrotic therapy to maximize clinical benefit and potentially mitigate downstream healthcare utilization ^2, 13^.

Despite the clinical and economic importance of antifibrotic therapies, there is a paucity of data on recent trends in Medicare Part D spending, utilization, and cost-per-claim for nintedanib and pirfenidone. Previous research has focused on other pulmonary medications or on aggregate drug spending, leaving a gap in the literature regarding antifibrotic agents. Understanding these trends is essential for informing policy decisions, optimizing resource allocation, and ensuring equitable access to evidence-based therapies. This study aims to address this gap by providing a comprehensive analysis of Medicare Part D expenditure and utilization patterns for antifibrotic therapies from 2019 to 2023.

## Methods

A retrospective, descriptive analysis was conducted using the publicly available Medicare Part D Drug Spending Dashboard data for the years 2019 through 2023. Annual data for nintedanib (Ofev) and pirfenidone (Esbriet) were extracted, including total gross spending, number of claims, and average spending per claim. Data were aggregated at the national level, and trends were assessed over the five-year period. No patient-level data were accessed, and all analyses were performed using summary-level information. The study focused on identifying changes in total expenditure, utilization patterns, and cost-per-claim for each antifibrotic agent. This approach aligns with prior pharmacoepidemiologic studies evaluating the adoption and cost trends of antifibrotic therapies in the United States ^9, 10, 13^.

## Results

From 2019 to 2023, total Medicare Part D spending on nintedanib (Ofev) increased from $789 million to over $1.8 billion. During the same period, spending on pirfenidone (Esbriet) declined from $617 million to $93 million. The number of claims for nintedanib rose from 77,655 in 2019 to 140,681 in 2023, while claims for pirfenidone dropped sharply from 65,773 to 9,204.

The average cost per claim for nintedanib increased from $10,167 in 2019 to $13,058 in 2023. For pirfenidone, the average cost per claim also rose, peaking at over $10,000 in 2023 despite the marked decline in utilization.

These data demonstrate a shift in Medicare Part D antifibrotic therapy expenditures, with nintedanib now accounting for the majority of spending and utilization. The observed divergence in Medicare Part D spending on nintedanib and pirfenidone from 2019 to 2023 reflects evolving clinical practice, market dynamics, and the broader context of antifibrotic therapy in IPF and PF-ILDs.

## Discussion

The marked increase in nintedanib spending and claims, alongside the sharp decline in pirfenidone utilization, may be hypothesized to result from several factors. These include differences in tolerability profiles, prescriber familiarity, patient preferences, and possibly formulary or insurance coverage changes ^9, 10^.

Nintedanib is more widely utilized than pirfenidone in the treatment of IPF and PF-ILDs within the Medicare population for several interrelated reasons. Nintedanib has received FDA approval for a broader range of indications, including not only IPF but also systemic sclerosis-associated ILD (SENSCIS trial) and PF-ILDs of diverse etiologies (INBUILD trial), whereas pirfenidone is only approved for IPF in the U.S ^14, 15^. This broader label allows nintedanib to be prescribed for a wider patient population and may facilitate insurance coverage and prior authorization for non-IPF fibrosing ILDs ^14^. The INBUILD and SENSCIS trials demonstrated that nintedanib significantly slows FVC decline in both PF-ILD and systemic sclerosis-ILD, respectively, with consistent benefit across subgroups, supporting its use in a range of fibrosing ILDs ^15, 16^. These robust data have influenced both clinical and payer preferences, as reflected in the American Thoracic Society guideline recommendations ^2^.

Studies indicate that while both drugs are effective, nintedanib may be associated with higher rates of gastrointestinal adverse events, whereas pirfenidone is more often linked to photosensitivity and rash ^7, 8^. Discontinuation rates due to adverse events are similar, but persistence may be longer with pirfenidone in some cohorts ^10, 11^. The lack of head-to-head randomized trials and the absence of a clear guideline preference for one agent over the other underscore the need for individualized treatment decisions ^2^.

The rising average cost per claim for both agents, despite declining pirfenidone use, highlights the broader challenge of specialty drug pricing in Medicare Part D ^17, 18^.This trend is consistent with national data showing that price increases, rather than utilization, are the primary driver of rising drug expenditures in Medicare ^17, 18^. The economic burden of antifibrotic therapy is substantial, with annual drug costs exceeding $70,000 per patient in some analyses, and out-of-pocket costs remaining high for beneficiaries ^9, 10^. While antifibrotic therapy is associated with reduced mortality and hospitalization rates among Medicare beneficiaries, the cost-effectiveness of these agents remains debated, particularly in the context of limited survival benefit and persistent disease progression ^10–13^.

From a policy perspective, these findings have several implications. First, the dominance of nintedanib in Medicare spending may prompt payers and policymakers to re-evaluate formulary management, negotiate pricing, and consider value-based reimbursement models ^17,18^. Second, the high and rising costs of antifibrotic therapy may exacerbate disparities in access, particularly for patients with limited financial resources or high out-of-pocket obligations ^17,18^. Third, the lack of generic alternatives and the slow uptake of biosimilars in this therapeutic class may limit opportunities for cost containment ^17,18^. Further evaluation is warranted to elucidate the drivers of shifting prescribing patterns, assess the comparative effectiveness and safety of nintedanib and pirfenidone in diverse populations, and explore the impact of policy interventions on drug utilization and patient outcomes ^7, 8^. Future research should also address the long-term cost-effectiveness of antifibrotic therapy, the role of patient-reported outcomes in treatment selection, and the potential for novel agents to improve clinical and economic outcomes in fibrotic lung disease ^4, 5, 15^.

In the context of escalating drug costs, recent health policy initiatives such as the Inflation Reduction Act (IRA) of 2022 have aimed to curb Medicare Part D spending through price negotiations for select high-expenditure medications, with the Centers for Medicare & Medicaid Services (CMS) empowered to negotiate prices for a limited number of high-spending Part D drugs each year, beginning with 10 drugs in 2026 and expanding to 60 by 2029 ^19^. Although antifibrotics like nintedanib and pirfenidone have not yet been targeted, their cost profiles suggest they could be included in future rounds of negotiations ^20^. If selected, price caps or reimbursement restructuring could significantly impact both Medicare expenditure and patient access.

Despite increasing spending, neither pirfenidone nor nintedanib has faced meaningful generic competition in the U.S. as of 2025. Pirfenidone, particularly, was excluded from Medicare price negotiation due to its status as a sole orphan drug, which confers extended exclusivity and delays generic entry. Such sole orphan drugs generate considerable revenue in the absence of generic competition, which contributes to sustained high prices and significant Medicare spending ^21^. Looking ahead, emerging antifibrotics may alter the treatment landscape. If approved, these novel agents could introduce competition, potentially reshaping prescribing trends and expenditure distribution within the Medicare system ^22^.

### Limitations

This study has several limitations. First, it relies on aggregated Medicare Part D data, which lacks individual-level detail and clinical outcomes. As such, the dataset cannot capture patient demographics, comorbidities, or drug adherence, limiting the scope of inference. Second, the findings are not generalizable to populations covered under Medicaid, commercial insurance, or uninsured individuals, who may exhibit different cost or prescribing patterns. Third, the reported drug costs are gross values and do not reflect manufacturer rebates or net expenditures, which are often negotiated privately and remain undisclosed. Fourth, this analysis cannot determine the exact clinical or administrative drivers behind observed shifts in utilization, such as adverse effect profiles, guideline changes, or market access issues. Lastly, the lack of inflation-adjusted dollar values may overstate real growth trends over the 5-year span.

## Conclusion

In summary, the current Medicare Part D expenditure landscape for antifibrotic therapies is shaped by clinical efficacy, tolerability, market forces, and policy constraints. Ongoing surveillance and research are essential to optimize the value and accessibility of these life-prolonging therapies for patients with IPF and PF-ILDs.

## Data Availability

The data underlying the results presented in this study are publicly available from the Centers for Medicare & Medicaid Services (CMS) Medicare Part D Drug Spending Dashboard: https://data.cms.gov/summary-statistics-on-use-and-payments/medicare-medicaid-spending-by-drug/medicare-part-d-spending-by-drug/data

## Acknowledgments

The author thanks Dr. Anthony Saleh of the Division of Pulmonary and Critical Care Medicine at NewYork-Presbyterian Brooklyn Methodist Hospital for his guidance and feedback during the development of this manuscript.

---

**Figure 1–3.**
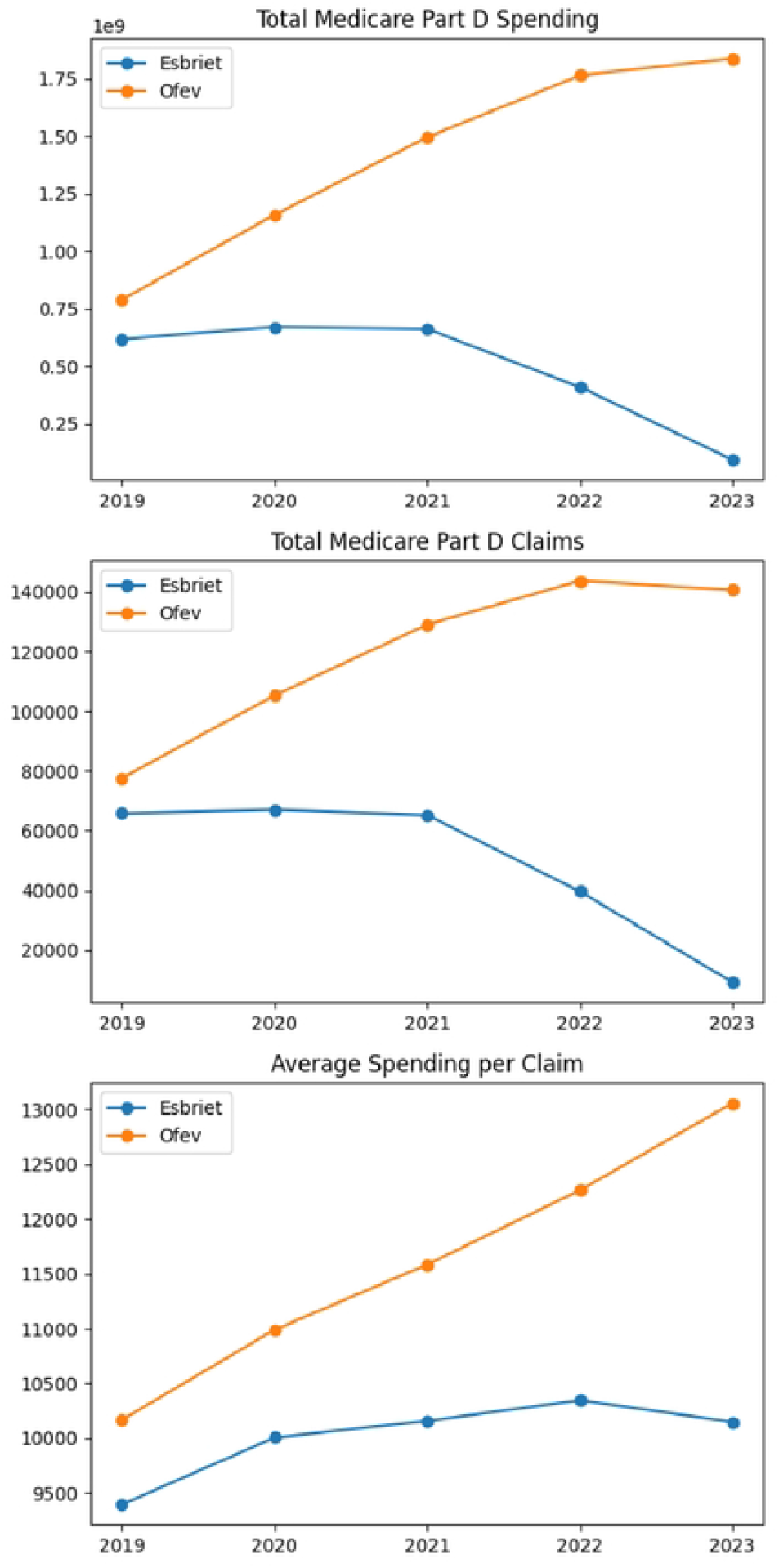
Trends in Medicare Part D spending, claims, and average spending per claim for Esbriet and Ofev (2019–2023).

**Table 1.**
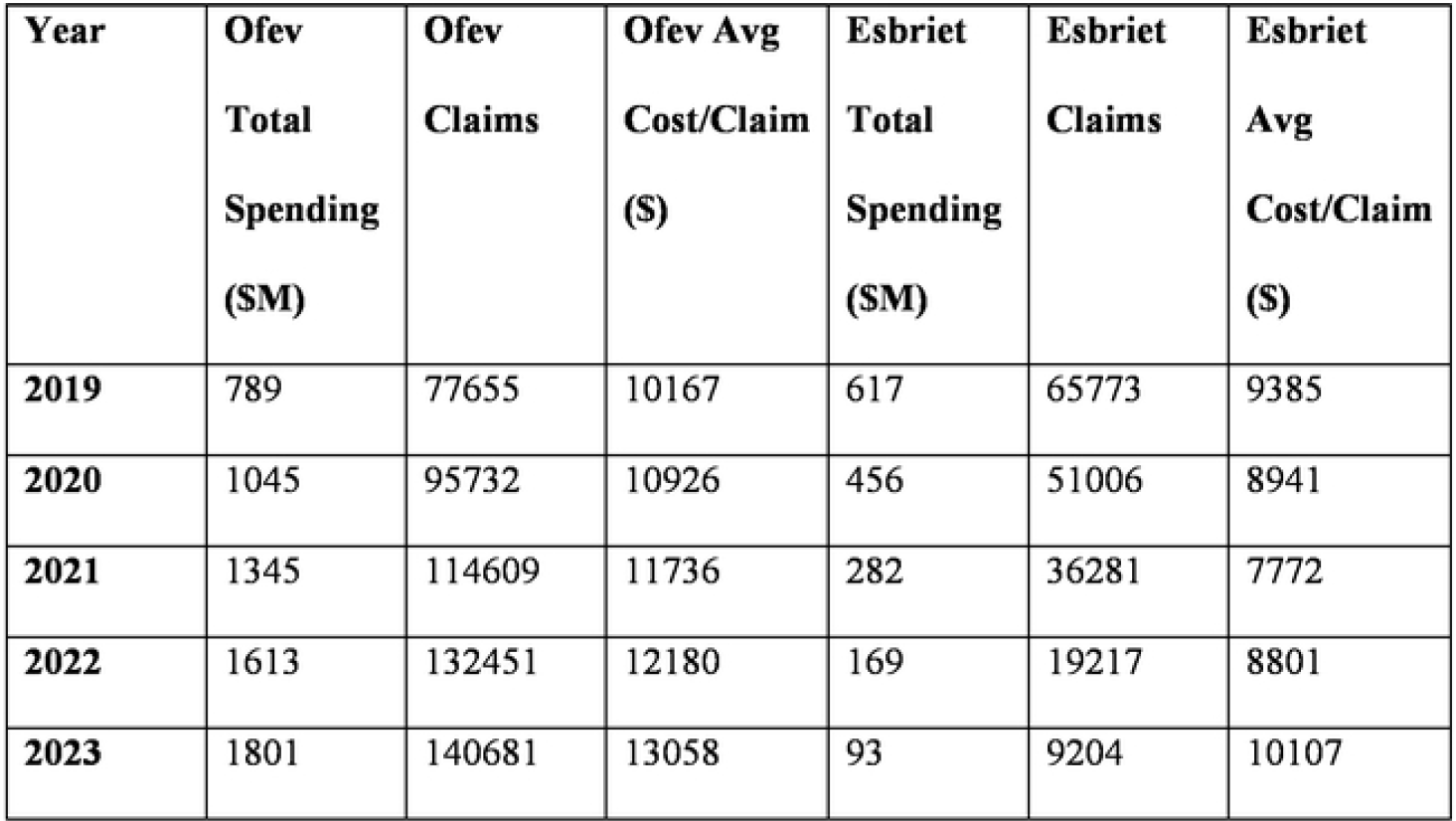
Medicare Part D Antifibrotic Drug Utilization and Spending (2019-2023)

